# “*We don’t routinely check vaccination background in adults”*: A national qualitative study of barriers and facilitators to vaccine delivery and uptake in adult migrants through UK primary care

**DOI:** 10.1101/2022.03.11.22272274

**Authors:** Jessica Carter, Anushka Mehrotra, Felicity Knights, Anna Deal, Alison F Crawshaw, Yasmin Farah, Lucy P Goldsmith, Fatima Wurie, Yusuf Ciftci, Azeem Majeed, Sally Hargreaves

## Abstract

**Background:** The COVID-19 pandemic has highlighted shortfalls in the delivery of vaccine programmes to some adult migrant groups; however, little is known around care pathways and engagement of these older cohorts in routine vaccinations in primary care, including catch-up programmes. Guidelines exist, but the extent to which they are put into practice and prioritised is unclear.

**Objectives:** To explore the views of primary care professionals around barriers and facilitators to catch-up vaccination in adult migrants (defined as foreign born; over 18 years) with incomplete or uncertain vaccination status and for routine vaccines to inform development of future interventions to improve vaccine uptake in this group and improve coverage.

**Design:** Qualitative interview study with purposive sampling and thematic analysis

**Setting:** UK primary care, 50 included practices.

**Participants:** 64 primary care professionals (PCPs): 48 clinical including GPs, Practice Nurses and healthcare assistants (HCAs); 16 administrative staff including practice managers and receptionists (mean age 45 years; 84.4% female; a range of ethnicities).

**Results:** Participants highlighted direct and indirect barriers to catch-up vaccines in adult migrants who may have missed vaccines as children, missed boosters, and not be aligned with the UK’s vaccine schedule, from both a personal and service-delivery level, with themes including: lack of training and knowledge of guidance around catch-up vaccination among staff; unclear or incomplete vaccine records; and lack of incentivization (including financial reimbursement) and dedicated time and care pathways. Adult migrants were reported as being excluded from many vaccination initiatives, most of which focus exclusively on children. Where delivery models existed they were diverse and fragmented but included a combination of opportunistic and proactive programmes. PCPs noted that migrants expressed to them a range of views around vaccines, from positivity to uncertainty, to refusal, with specific nationality groups reported as more hesitant to get vaccinated with specific vaccines, including MMR.

**Conclusions:** WHO’s new Immunization Agenda (IA2030) has called for greater focus to be placed on delivering vaccination across the life-course, targeting under-immunised groups for catch-up vaccination at any age, with UK primary care services therefore having a key role to play. Vaccine uptake in adult migrants could be improved through implementing new financial incentives or inclusion of adult migrant vaccination targets in QOF, strengthening care pathways and training, and working directly with local community groups to improve understanding around the benefits of vaccination at all ages.

## Introduction

Adult migrants in Europe – particularly those from low- and middle-income countries – may be at risk of under-immunisation for routine vaccinations due to missed vaccines and doses as children (due to lack of availability, war/disruption, poorly functioning health systems, and personal, social, and physical barriers to accessing vaccines), and/or missed boosters, and differing vaccination schedules in their home country (especially for newer vaccines such as HPV), and so may not be aligned with the UK’s vaccination schedule on arrival (1, 2). Additional vaccines may be recommended if they return to their home country, or for specific occupations (eg, tetanus and hepatitis B). Some migrant populations are known to be at risk of under-immunisation (2-5) and were involved in recent outbreaks of vaccine-preventable diseases in Europe, including measles (1). However, adolescent and adult migrants, beyond school age, are often not routinely incorporated into vaccination programmes on arrival to most European countries, including the UK (6). The COVID-19 pandemic has highlighted shortfalls in engaging older migrants, and other marginalised groups, in vaccination programmes (7), yet it has also presented a range of new opportunities and innovations in vaccine service delivery and policy making to these groups, which merit greater consideration beyond the pandemic.

The World Health Organization’s new Immunization Agenda 2030 (IA2030) (8) aims to improve vaccine coverage for vaccine-preventable diseases, placing an emphasis on achieving equitable access for vulnerable populations and integrating catch-up vaccination for missed vaccines and doses throughout the life-course. WHO recommends that all countries have a catch-up vaccination policy and catch-up vaccination schedule in place, to close immunization gaps that would otherwise compound as populations increase in age (9, 10), and that it is always “better to vaccinate late than never”. Although age limits apply for administration of a small number of vaccines, for most VPDs, providing vaccines late will still result in protection against morbidity and mortality, as well as reducing transmission and risk of outbreaks, with personal and community-level benefits. Specific WHO guidance for catch-up vaccination is available (9); in Europe the European Centre for Disease Prevention and Control (ECDC) has published guidance on catch-up vaccination in children and adult migrants on arrival (11, 12), calling for healthcare providers to consider revaccinating adult migrants with uncertain vaccination status or no recorded history of vaccination. For UK arrivals, advice is available from the UKHSA on the ‘vaccination of individuals with uncertain or incomplete immunisation status’ (see Panel 1), which will be relevant to most arriving migrants (13). However, the extent to which these guidelines and policies are put into practice and prioritised by UK primary care – tasked with delivering the majority of the UK’s vaccine programme – is not known. No studies to date have explored the views and experiences of front-line primary care teams on approaches to catch-up adult vaccination in arriving migrants. We therefore did a national qualitative in-depth interview study with a range of primary care professionals to understand the challenges and needs of migrant populations with regards to catch-up vaccinations programmes, and facilitators and solutions to addressing gaps in service provision.

### Panel 1

**Vaccination of individuals with uncertain or incomplete immunisation status. Reproduced from (13)**.

From tenth birthday onwards:

- Td/IPV and MenACWY*and MMR *Four week gap* Td/IPV and MMR *Four week gap* Td/IPV

First booster of Td/IPV – preferably 5 years following completion of primary course. Second booster of Td/IPV - ideally 10 years (minimum 5 years) following first booster.

- HPV:

- all females who have been eligible remain so up to their 25th birthday
- males born on or after 01/09/2006 are eligible up to their 25th birthday

- Subsequent vaccination – as per UK schedule (see Flu Vaccine, Shingles vaccine, PPV) and COVID-19.

*\*Those aged from 10 years up to 25 years who have never received a MenC-containing vaccine should be offered MenACWY. Those aged 10 years up to 25 years may be eligible or may shortly become eligible for MenACWY usually given around 14 years of age. Those born on/after 01/09/1996 remain eligible for MenACWY until their 25th birthday*.

## Methods

### Design

Qualitative semi-structured interviews of both clinical and administrative staff were undertaken by telephone, following a topic guide collaboratively developed by the research team with support from a board of migrant representatives. The guide was piloted prior to data collection and iteratively developed throughout the data collection process, with the addition of further prompts and probes to develop richer understanding and addressed key areas around approach to vaccination of adult migrants, factors affecting vaccine hesitancy and uptake, and possible interventions to strengthen delivery (Panel 2). Ethics approval was granted by the Health Research Authority (reference number: REC 20/HRA/1674).

#### Panel 2

**Topic guide**

Background Questions:

- Proportion of migrants seen at practice, migrant health training and experience
- General barriers and facilitators to registration and provision of care for migrant patients

Questions pertaining to Vaccination of Adult Migrants:

- Are you aware of any guidance regarding vaccination and infectious disease screening in migrants?
- Have there been any outbreaks of vaccine preventable diseases or cases of vaccine preventable diseases in your area involving migrants – we are particularly interested in adults? (If yes, what do they think the reasons might be?
- What experience have you had with adult migrant patients and vaccination?

Questions regarding Practice Approach to Vaccination of Adult Migrants:

- How do you approach catch-up vaccination in the adult migrant patient group, specifically ensuring adult migrants are caught up to align with the UK schedule?
- Who is responsible for vaccination at your practice?
- Is there a mechanism at your practice or in your area to engage adult migrants on catch-up vaccination?
- Is there a local catch-up vaccination pathway?
- Do you target any specific groups?

Questions regarding possible interventions to increase uptake of catch-up vaccination in migrants:

- If there are no mechanisms/pathways in place locally do you think there should be?
- What could such a system look like?
- Are you aware of any other interventions relating to vaccination in migrants? If so, what made them successful/ unsuccessful?
- What do you think about a migrant health check, and what vaccinations would be important to cover in this for adult migrants in your view?
- What in your opinion would be the key to a successful intervention/ behaviour change in primary care?

### Setting

Participants were recruited from 50 GP practices in urban, suburban, and rural settings across England. Practices were based in one of six local clinical research networks (CRNs) — CRN Kent, Surrey and Sussex; CRN South London; CRN North Thames; CRN North West London; CRN West Midlands; and CRN Greater Manchester with the exception of a practice in Newcastle and another in Oxford.

### Participants

Participants were purposively sampled to capture the diversity of experiences in general practice, from administrative and clinical primary care roles, and practices which varied both in size, and urban/rural location, factors which could influence the number of migrant patients and the organisation of care. Recruitment occurred via local Clinical Research Networks, ‘word of mouth’ invitations from colleagues and a number of primary care newsletters, social media groups and practice manager mailing lists. All participants who expressed an interest in taking part were e-mailed a participant information sheet and consent form and invited to a telephone interview at a time of mutual convenience, with written informed consent being given in advance. £20 vouchers were given as compensation for each participant’s time.

### Data collection

Telephone interviews, between 30-60 minutes, were carried out by JC (GP) FK, (GP registrar) and AD and AFC (academic researchers). Findings from the initial interviews were discussed across the group and led to the development of additional prompts and lines of questioning in the topic guide, as well as additional lines of questioning for non-clinical participants. All but three of the interviews were digitally recorded and transcribed verbatim. The remaining three were lost through technical error but were typed up from extensive field notes. Transcripts were anonymised with a coded participant number and checked for accuracy. Data collection continued until there was thematic saturation (14) across all core themes as unanimously agreed across the team.

## Data Analysis

Data analysis was inductive, based on the stages of thematic analysis (15). The transcripts were read repeatedly by AM (familiarisation) and emerging themes and patterns were identified and discussed with FK and JC who had also previously immersed themselves in the data. Initially, a coding of 10 transcripts on Microsoft Excel by AM allowed identification of emergent themes and discussion with FK and JC. NVivo (version 13) was then used to organise codes and iteratively refine and develop the emerging coding framework through a process of constant comparison, with close attention paid to non-confirmatory cases which contradicted existing themes. The final coding and themes were conceptualised through recurrent discussion by AM, FK, JC and SH. Active reflexivity was attempted from the study’s onset, and input from across the multidisciplinary team, with support from the migrant advisory board, facilitated robust discussion throughout.

## Results

In total, 64 interviews were conducted. 48 interviews were held with primary care staff: 25 GPs, 15 practice nurses, seven healthcare assistants (HCAs), one clinical pharmacist, 11 practice managers, and five receptionists. The majority of staff (50 [78.1%]) worked in urban practices. Characteristics of included participants are presented in Table 1.

**Table 1:** Characteristics of included participants.

| Characteristics | Total participants (n=64) |
| --- | --- |
| Staff type | General Practitioners (GPs): 25 (39%)<br>Practice Nurses (PNs): 15 (23.5%)<br>Healthcare assistants (HCAs): 7 (11%)<br>Pharmacist: n=1<br>Practice Managers: 11 (17%)<br>Receptionists: 5 (8%) |
| Ethnicity | African: 4 (6.3%)<br>Other Asian background: 2 (3.1%)<br>Mixed: 3 (4.7%)<br>Other white: 5 (7.8%)<br>Caribbean: 1 (1.6%)<br>Indian: 11 (17.2%)<br>Pakistani: 3 (4.7%)<br>White British: 32 (50%)<br>White Irish: 2 (4.7%) |
| Age | 45 years (SD 11.8 years) |
| Sex | Female: 54 (84.4%)<br>Male: 10 (15.6%) |

### Barriers reported by PCPs to vaccine uptake in adult migrants

Participants reported that their migrant patients express a range of views around vaccines from positivity to uncertainty, to refusal. They highlighted direct and indirect barriers to vaccine uptake at both patient and staff level, as well as specific issues relating to many migrant groups including specific nationalities. Generalised mistrust and misinformation about vaccinations in migrant groups was commonly reported, which was often perceived by PCPs as resistance to information-sharing about the vaccine in question.

*“It’s really hard to break through that barrier of… this is the evidence [about this vaccine]… I don’t think they’re listening… they’re thinking… this is someone from my community saying this [other information]. And you’re not from my community… I don’t know if you have the best interests [in mind]*.*” GP10*

Some PCPs gave their views on vaccine acceptance and uptake linked to specific nationalities, and most often reported beliefs or experiences that migrants originating from Eastern Europe, France and Italy, Somalia and Bangladesh tend to be hesitant about vaccines. Fixed negative views around vaccines were most often reported from Eastern European migrants, who were also viewed as having poor vaccination records and as wanting to follow a different vaccination schedule (as per protocols in their home country), with some returning to their own countries to be vaccinated. (Table 2). The doctor-patient relationship was highlighted as a key factor in tackling mistrust and vaccine hesitancy; some PCPs felt it was easier for migrant patients to connect with PCPs from their own communities.

**Table 2:** Perceptions of staff around acceptance and uptake in specific nationality groups.

| Participant reporting | Quote |
| --- | --- |
| GP | “Yes, I think the MMR would be a good example, where...I can remember a conversation I had with a mum whose friend, who was also Bangladeshi... Their child was off sick and they blamed it on the MMR vaccination, because before the vaccination he was fine...” |
| GP | ...now it’s more Bangladeshi, so Somalian was really with the MMR thing. But we still find more Bangladeshi families delaying or refusing the immunisation of their babies.....So, yes they always blame... This is too much, the baby is young, we’re not sure about the long term effects. |
| GP | “[The Somalian population]...is a massive concern for us, with regards the patients unfortunately, falsely attributing MMR with an autism link” .....” I think it was the belief of autism, but why more in the Somali community than any other minority group, I’m not too sure.” |
| HCA | [The Somalian population are] ... very happy to vaccinate as elderly patients. But, [they think]...the children will get something, get over it. And I think with MMR, they do feel that there's side effects. They think that it causes Autism and things like that. |
| Practice nurse | I don't know where, Somalia or Eritrea that there was only one interpreter in London who could speak their language. Even their care worker obviously could not speak their language. And so, trying to get immunisation history or any history out of these two young men was totally impossible |
| HCA | I would say that Europeans [migrants], they refuse because they think they've had them, even if it's been a long time and they don't know. |
| GP | What I have noticed is that when a patient comes from... Eastern European countries... they do come in with a vaccination record. It's usually incomplete... and sometimes we doubt [it is true and], whether...you can pay someone to give you a vaccination record but it actually hasn't happened. |

Indirect barriers, such as mistrust of the NHS system generally and inability to communicate vaccination histories, were felt by participants to reduce the likelihood of migrants accessing catch-up vaccinations. Issues were raised about immigration status and fears about being reported to authorities, and language barriers, including lack of written communication about vaccine services.

*“I think immigration status, out of anything, is going to be the main issue. A lot of people that live in this country without status, going to the GP is a massive risk*.*” PN 13*

*“Language can be a barrier for subtleties of communication, despite language line” GP21*

*“I think we probably ought to translate that communication [about vaccine programmes] in written Bengali, and perhaps Somali as well*.*” GP10*

*“There’s usually a long wait and possibly a language barrier as well that may stop [people] from communicating or trying to make that appointment” PN 15*

Direct health-system barriers to providing catch-up vaccination for adult migrants included lack of training among staff and lack knowledge of guidance around catch-up vaccination. In addition, participants raised the fact that unclear or poorly documented vaccination histories meant staff were unclear as to what to do, as well as highlighting problems with vaccination records not being transferred within the NHS, and a lack of availability of records from migrants’ home countries, including limited translation of previous records into English. Some migrants were reported as having different ages recorded, leading to challenges determining vaccine eligibility.

*“And we’re certainly not being given any records from other countries that might support [vaccination catch-up]… unless the patient is super well-organised and providing that it happens to be in English or a language that’s directly transferrable…” Admin 6*

*“The nurses would need some kind of education in how to complete incomplete vaccination programmes in adults” Admin 12*

*“So, no, I’m not aware of any guidance for [vaccination in] migrant people” GP 2*

There were also a number of additional barriers to accessing care at staff and system level which were felt to indirectly reduce the likelihood of adult migrant patients being offered and accepting a catch-up vaccine or travel vaccines through the travel clinic. These included a lack of time to carry out proactive catch-up programmes, or to follow up on opportunistic or challenging conversations where a vaccination need was highlighted, especially when using a translator. The financial pressures and impact of vaccination programmes falling outside of current incentive schemes, such as QOF, also impacted on the time available for the programmes.

*“It’s just time pressure, the way that the general practice is working at the moment unfortunately is reactive…And so, with things like vaccinations, especially if it’s catch up or screening, can always wait… [because] you’re going to deal with [someone’s chest infection or…diabetes] before you deal with their symptomatic screening. GP6*

*There are “no incentives for catchup vaccination, MMR… especially compared to childhood immunisations and chronic diseases in QOF*.*” (GP16)*

Multiple barriers were identified in relation to specific vaccines in the UK schedule, with a summary of key themes, by vaccine, reported in Table 3.

**Table 3:**
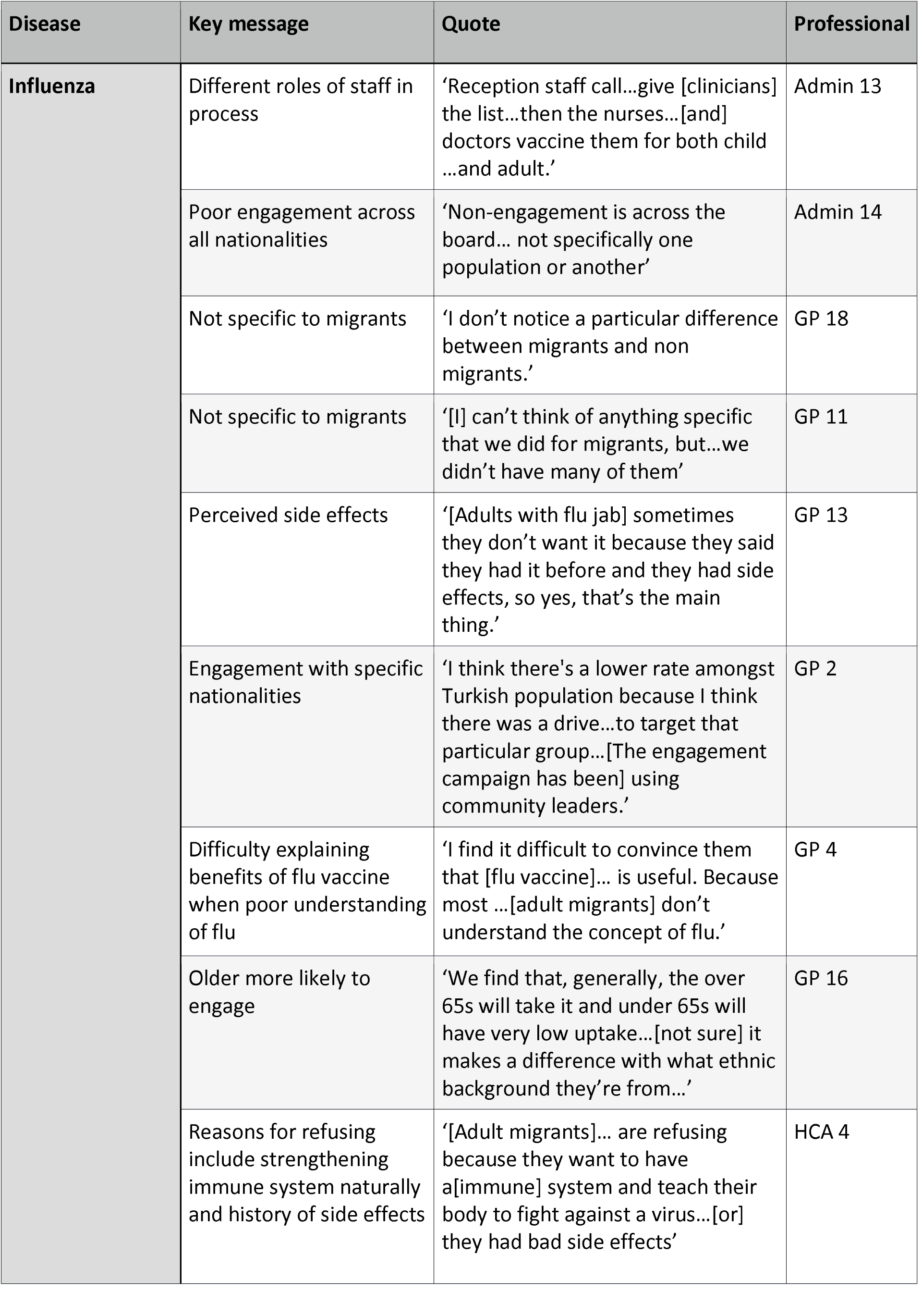
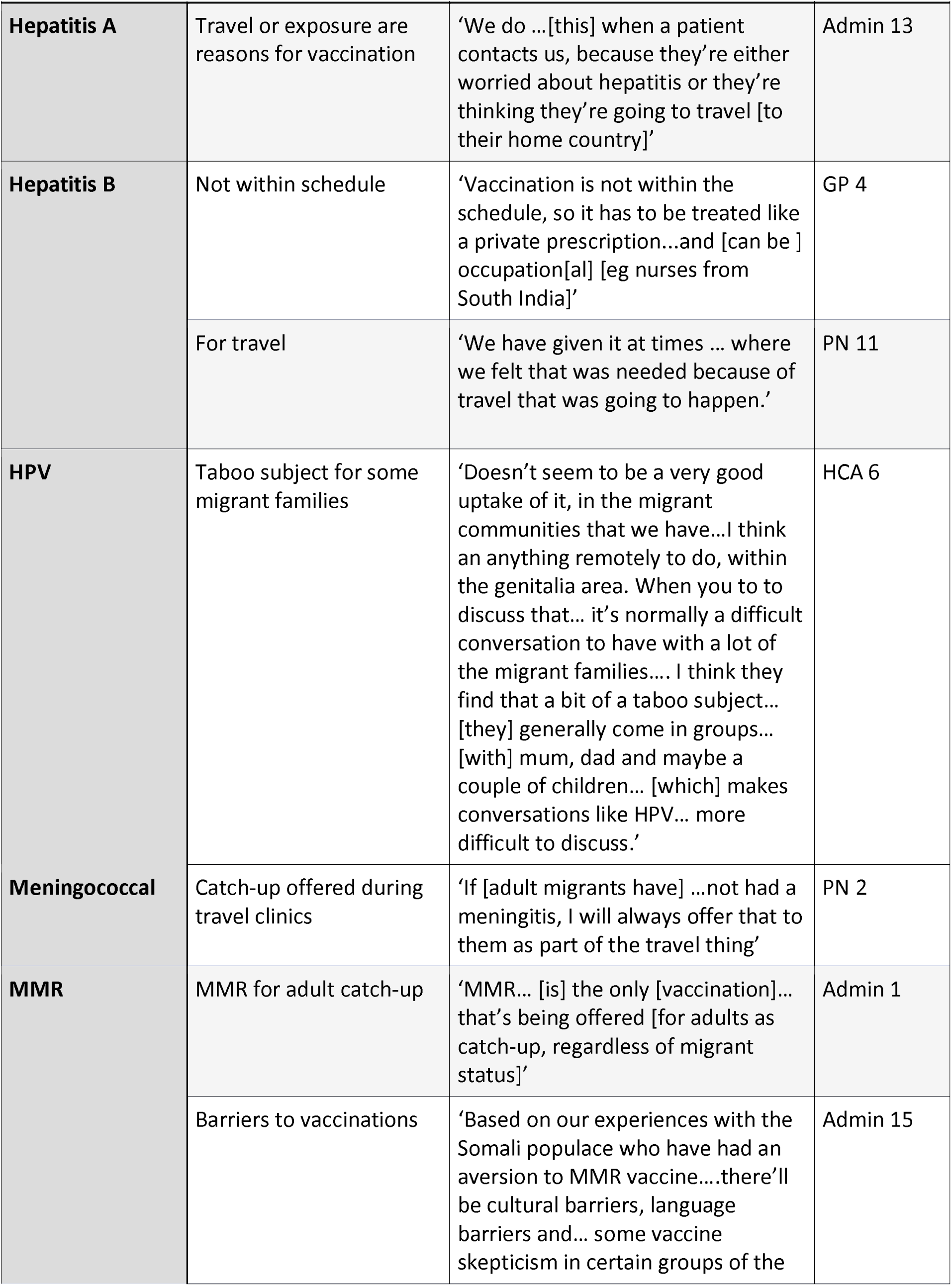
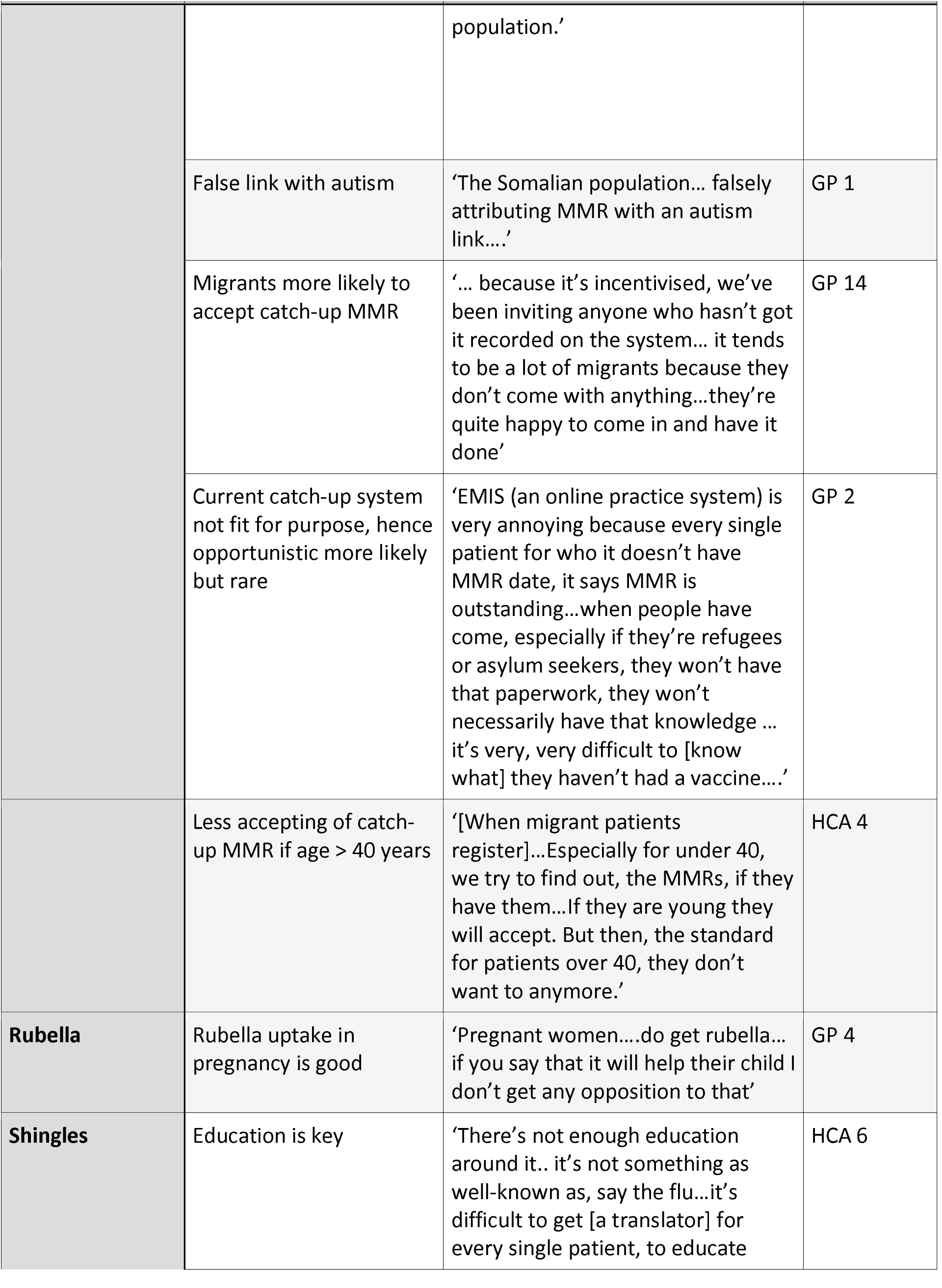
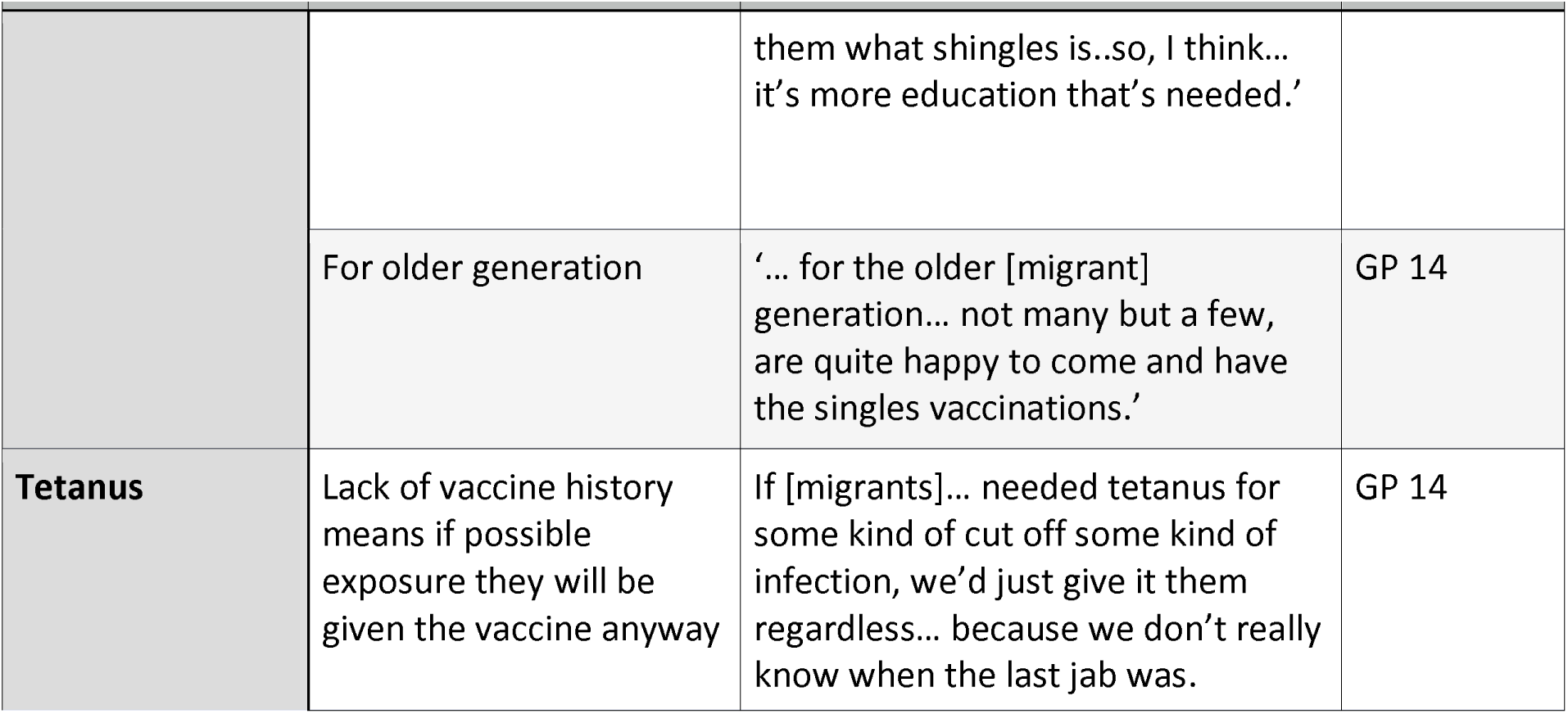
Key issues pertaining to adult migrants for specific vaccines in the UK schedule.

### Fragmented models for vaccine delivery to adult migrants

Almost all clinical staff reported the availability of good catch-up programmes for childhood vaccination among recently arrived migrants, with some PNs specifically quoting the Public Health England Schedule for individuals with uncertain vaccination status. Incentivization for under-5s vaccination included the Quality Outcome Framework (QOF), and well-resourced systems to ensure children are not missed, including the vaccination record ‘red book’ and using recall systems to contact patients, such as sending reminder texts. By contrast, adult migrants were often reported as being excluded from vaccination initiatives. One GP stated that over 5’s and adults are sometimes assumed to be *“up to date from the country they come from”*, and many staff, especially GPs and administrators, were not aware of any catch-up vaccination programmes for adult migrants.

*“We don’t routinely check vaccination background in adults” GP 16*

*“We do catch-up vaccinations for children and young adults who’ve missed their primary vaccinations, but in terms of adults or people who are arriving to the UK, no” Admin 6*

*“Ad hoc. We haven’t had a particular programme for [adult catch up vaccinations] GP15*

Where adult catch-up vaccination was provided, models of delivery were diverse and fragmented, comprising a range of clinics and providers, different staff members (primarily nurses), and a combination of opportunistic and proactive programmes. Providers of catch-up vaccination for adult migrants included: NHS GP practices, detention centres (for undocumented migrants and asylum seekers), migrant-specific or language-specific clinics, private clinics and specialised clinics (e.g. sexual health clinic in China Town), with distinct benefits and challenges.

#### Detention centres

*“Interpreters weren’t always readily provided when I was at the detention centres. We found that really difficult and it took several visits [to determine which vaccines were required and these to be given]” PN13*

*“I can remember a conversation I had with a [Bangladeshi] mum…Their child was off sick and they blamed it on the MMR vaccination…[she] believed her friend over me… they’re thinking, … you’re not form my community… I don’t know if you have… my child’s best interests*.*” Whereas the doctor running the Bangladeshi clinic had more trust from their patients “Because she has contact with them regularly…they tend to connect with her very easily, because they feel … this doctor …[makes] the effort to get to know us, by virtue of just doing her clinic every week” GP 10*

Respondents reported vaccinations programmes were a mix of opportunistic and proactive delivery approaches. Proactive programmes included methods such as setting up searches, call and recall systems to contact patients, and targeted campaigns for specific vaccinations (eg flu).

*“We run recalls [for adult migrant catch-up] constantly throughout the year. We will target separate cohorts of patients, just so we can make sure we’re recalling everybody”. Admin 5*

Opportunistic usually meant identifying a patient needed a vaccination when they were attending the practice for another reasons. The vaccine could be given immediately, or the patient booked into an appointment at a later date.

*“…if I notice and if I remember or have time to mention it, then I encourage people to… [but] they’re usually coming with quite a few issues, and we’re using an interpreter… there’s a lot to cover…[hence no time to cover vaccination]. GP 18*

There are also diverse approaches to vaccine delivery between practices, with different staff involved in different aspects of the vaccine programme. However, many programmes are nurse-led, with the practice nurse having main responsibility.

*‘[It’s a] mixture of me, one of the partners, and then the reception staff are the ones who actually call the patients and arrange for them to come in” Admin 13*

*“If they’re struggling to get somebody to agree [to take a specific vaccine]… we get the named GP*.. *to take responsibility for having that conversation and trying to talk them round” Admin 9*

*“…our vaccines are really well-run at the practice by one of the nurses in particular. She runs the whole immunisation program, the childrens, the flu, the catch up, everything. So, I would imagine that there’s probably a lot going on that I’m not aware of. I suspect and she always goes on updates and is very much aware of new guidance to things so I’m sure that she’s probably doing a lot of stuff behind the scenes that I’m not aware of*.*” GP 3*

### Travel Vaccination and Occupational vaccines

Provision of catch-up vaccines and additional vaccines to adult migrants was also mentioned in the context of travel and occupational requirements. Delivery of travel vaccinations was highlighted by a variety of participants for migrants visiting their home countries and travelling to Haj.

*“I think people are very good at knowing they need vaccinations, especially people who have been settled in England for quite a long time and are maybe making an infrequent return visit home to may visit relatives or family elders or to go for a celebration” Admin 6*

*“They will go for the bare minimum of what is offered, or what they need to have as certificated. If they’re doing the pilgrim to the Haj, then they have to get the meningitis. If they… need yellow fever, they’ll get the yellow fever…or they just don’t have anything*.*” PN 2*

Different nationalities were reported as having varying levels of engagement with travel vaccine uptake. One PN reported Bangladeshi families travelling more being ‘more engaged’ than Middle Eastern people. Another reported Europeans as *‘more engaged with travel clinics than*…*people… from Pakistan, India, Bangladesh and African countries”. [PN2]* African patients were described as having a poorer uptake of travel clinics than Europeans *“people returning to DRC or Tanzania…their uptake is poorer than younger European people” PN2*.

Participants noted that travel clinics can also be an opportunity for opportunistic adult catch-up: *“The nurses who do the travel clinics are certainly very switched on to catch-up vaccines and will make sure everybody’s up to date with DTP and MenACWY, even if they’re not going to a country for which you need ACWY*.*” GP 17*

Travel vaccines were often given privately due to recommendations these should be done outside of the core contract, and this was primarily the case for adults but not children.

*“We do … Hepatitis A and then typhoid as part of the core contract. Anything else we direct patients to a private travel clinic”* GP 24

However, there was variability in provision, with one GP stating: *“We don’t charge for anything, including malaria pills” (GP 17). This would impact the “migrant population who are going backwards and forwards to their home countries [and] constitute quite a large percentage of patients that we see for travel clinics” (GP 17)*

Occupational vaccines were mentioned as sometimes being provided ‘outside the schedule’ for healthcare staff, such as nurses.

*“ [Hepatitis B] vaccination is not within the schedule, so it has to be treated like a private prescription…some of them are nurses …[and they] usually come from the South Indian population. Carers and nurses” GP 4*

*“We shouldn’t be seeing people wanting occupational health-related vaccination, but we do often get people asking for that” PN 1*

### Strengthening vaccine delivery in UK primary care

Primary care staff raised a range of potential solutions and action points to increasing vaccine uptake, especially in adult migrants, including addressing personal, societal, and physical barriers to vaccination systems through UK primary care alongside financial incentives to primary care to deliver adult catch-up vaccination. Barriers and potential solutions raised by participants are summarised in Table 4.

**Table 4:**
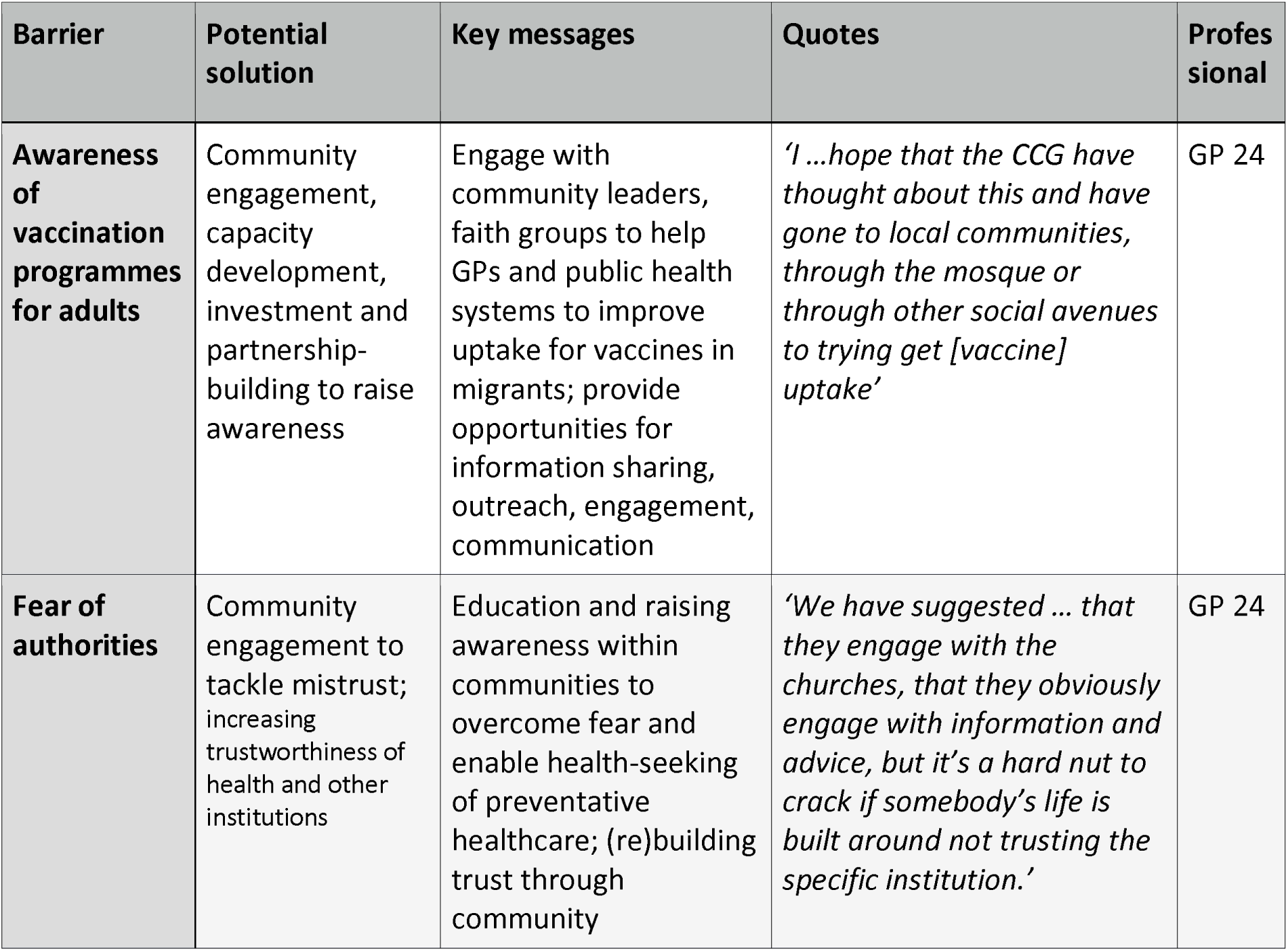
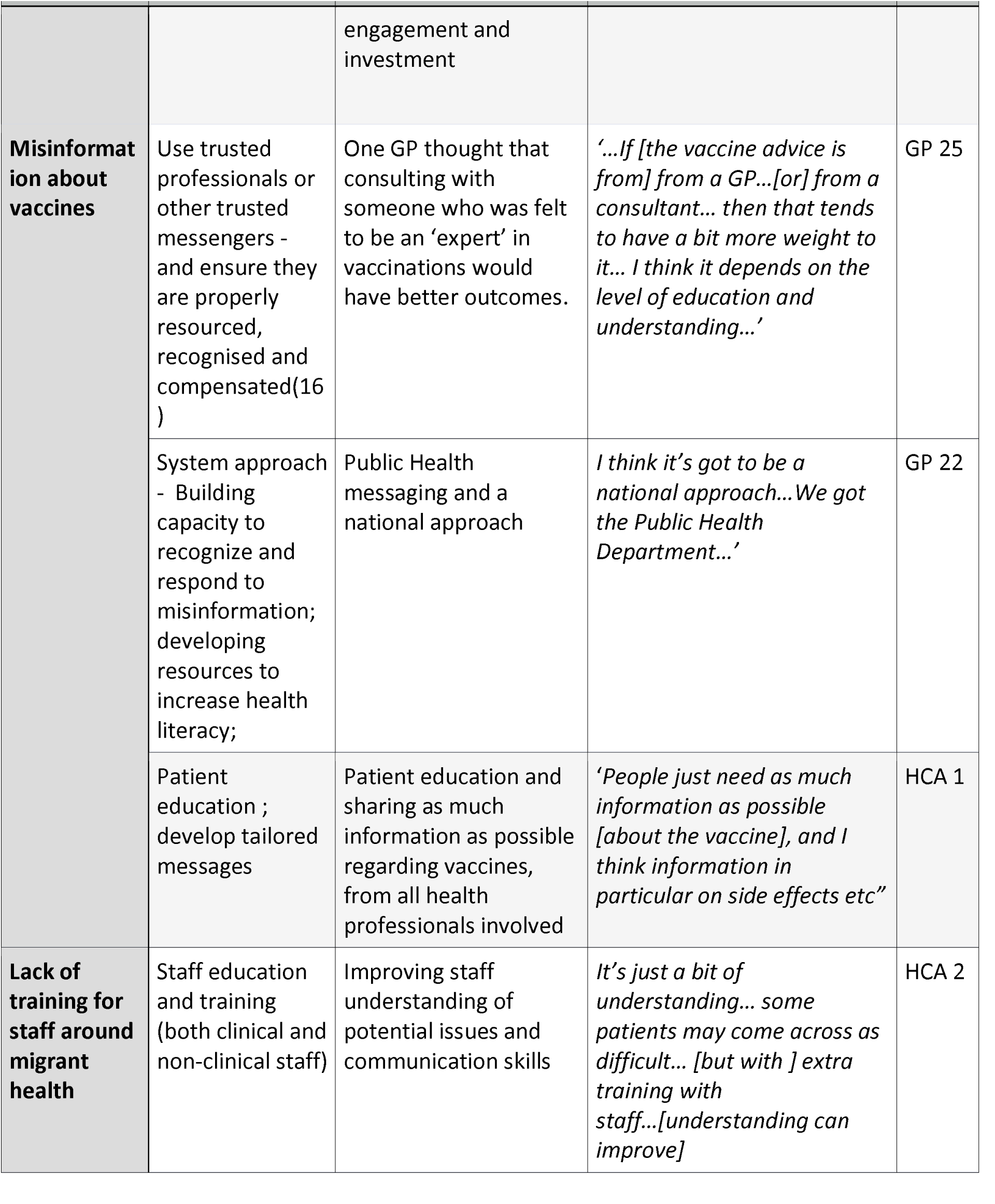
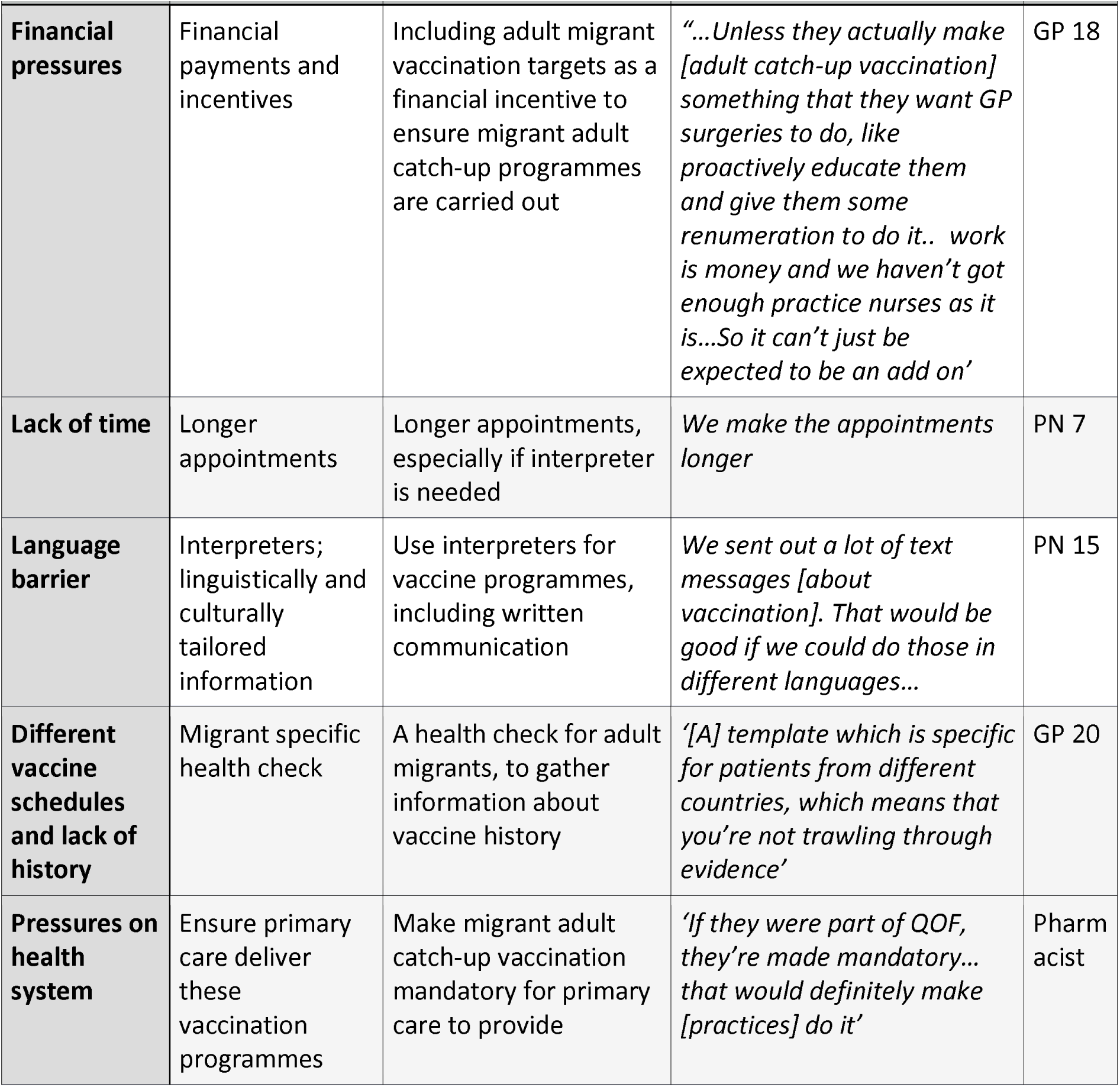
Barriers and solutions identified.

## Discussion

WHO’s new Immunization Agenda (IA2030)(8) has called for greater focus to be placed on delivering vaccination across the life-course, targeting under-immunised groups for catch-up vaccination at any age, with primary care services therefore having a key role to play in the UK context. In our study, however, participants highlighted direct and indirect barriers to delivering catch-up vaccines in adult migrants who may have missed vaccines as children, missed boosters, and not be aligned with the UK’s vaccine schedule. Barriers were noted at a personal and service-delivery level, with themes including: lack of training and knowledge of guidance around catch-up vaccination among staff; unclear or incomplete vaccine records; and lack of incentivization (including financial reimbursement), prioritisation, and dedicated time and care pathways. Adult migrants were therefore reported as being excluded from many vaccination initiatives, most of which focus exclusively on children. In addition, PCPs reported that migrant patients express a range of views around vaccines to them, from positivity to uncertainty, to refusal. Some migrants including Somali, Eastern-Europeans and Bangladeshi groups were often reported as being hesitant to get vaccinated, with specific concerns reported for specific vaccines, including MMR. Greater consideration needs to be placed on potential delivery points for catch-up vaccination in adult migrants – for example, local places of worship and other trusted or familiar sites – alongside offering financial incentives or inclusion of adult migrant vaccination targets in QOF. Improving uptake of catch-up vaccination in this group will require new care pathways and training of front-line staff, alongside working directly with local community groups to communicate the benefits of vaccination at all ages. In addition, greater collaboration across systems and community groups and culturally competent campaigns are warranted. At a time when COVID-19 vaccination programmes are being rolled-out across the world, this study adds important understanding regarding the specific vaccination needs and concerns of migrants, and the challenges faced by the staff delivering vaccination programmes to migrant populations and older cohorts.

A key strength of the study is the number and variety of primary care staff included from across England in diverse settings. Interviewees were a self-selecting group, which may have affected the profile of those responding – a common consideration in qualitative research. However, a breadth of practices were involved, including those that do not see many migrants, and this diversity and the scale of the study is likely to have added to the breadth of experience and solutions reflected in our findings, as well as enhancing the validity. We noted that often staff made broad generalisations about specific nationality groups, which needs to be considered when assessing findings. The structure and experience of primary care across Europe and between the devolved nations of the UK may differ so the recruitment only within England may limit the generalisability of the findings, however other European and international studies (6, 17, 18) have come to the similar conclusions in terms of healthcare provider, system, and patient-related barriers to catch-up vaccination in relation to adult migrants, so we feel that this would be unlikely.

We found a range of direct and indirect barriers to delivering catch-up vaccines in adult migrants who may have missed vaccines as children, missed boosters, and not be aligned with the UK’s vaccine schedule, from both a personal and service-delivery level. Our findings concur with those of similar study in Norway (17) which found no consistent or structured approach to vaccinating adult migrants in Norway, including no guidelines from governing bodies on how to organise vaccination to adult migrants. Reasons why adult vaccination is not prioritised included tuberculosis screening and treatment taking precedence, and a common assumption among healthcare providers that vaccinations are dealt with in childhood(17). A questionnaire survey of experts across Europe(6), and policy analysis(19), found that policies and practice differ across European countries with respect to adult vaccination and the inclusion of migrants in vaccine systems on arrival. Only 13 of 32 countries in the EU/EEA had policies in place to offer MMR vaccines to adult migrants, with 10 countries reporting that they would charge fees(6). Variations in vaccine policies targeting adult migrants were reported in another European survey (20). In addition, it is well known that some migrants face a range of barriers to health systems more broadly. This suggests that more inclusive policies are required placing an emphasis on new approaches to ensure older migrants are included, and that such policies are well implemented in practice.

Implementation will be key, and our study raised numerous points that merit greater consideration. Service delivery barriers have previously been described in other areas of migrant health, including screening for infection, with GPs citing concerns about lack of awareness around the health needs of migrants and insufficient time and resources (21, 22). It has previously been noted that negative biases from healthcare staff towards migrant patients or pre-conceptions about vaccine hesitancy in specific ethnic groups may have an impact on patient trust (23, 24), which is known to be a major factor in vaccine uptake (25). Education and training of front-line providers will be a critical component given the critical role that the PCP-patient relationship has for building trust in vaccination. This must involve raising awareness of the diverse experiences of migrants and how to approach potential vaccination concerns with sensitivity, as well ensuring an understanding around the potential the low vaccine coverage in their countries of origin as children, different dosing schedules, and particularly low coverage for newer vaccines. For HPV, for example, global coverage for the final dose was only 13% in 2021 (26) – suggesting many migrants aged under 25 years would be eligible for HPV vaccination as part of the UK’s more advanced programme. However, likely a key factor will be financial incentivisation to encourage practices to target potentially under-immunised adults for catch-up vaccines, which was a recurrent theme among those interviewed. Catch-up vaccination could be considered at various entry points, for example the New Patient Health Check or the NHS Health Check. Since April 2020, MMR now comes with an item of service payment, including for catch-up vaccination in patients who missed out on scheduled vaccines, which should encourage practices to offer appropriate vaccinations to patients regardless of age.

Tackling hesitancy and educating migrant and broader ethnic minority communities about the benefits of vaccination across the life-course will also be a critical component(22, 27), with COVID-19 presenting numerous innovations in service delivery in this area that merit further consideration to routine vaccination going forward including outreach, policy shifts to facilitate registration of migrants with primary care providers, and anonymous vaccination in trusted locations (22, 28). We found that certain nationality groups (Somali, Eastern-Europeans and Bangladeshi) may be more hesitant to receive vaccines than others, or reluctant to receive certain vaccines, aligning with a recent systematic review that found nationality/country of origin to be a key determinant of vaccine uptake for routine vaccines and COVID-19 vaccines in European datasets (7). In this study, acceptance barriers were mostly reported in Eastern European and Muslim migrants for HPV, measles, and influenza vaccines, with 23 significant determinants of under-vaccination in migrants found (p<0.05), including African origin, recent migration, and being a refugee/asylum seeker (7). A systematic review of interventions to improve vaccination uptake in newly-arrived migrants to the EU/EEA (29) highlighted the potential solutions of social mobilization and outreach programmes, planned vaccinations, and educational campaigns. Our data points to a recommendation for policy makers to include adult migrants specially in catch-up vaccination programmes on arrival, and to ensure policy around the delivery of catch-up vaccination across the life-course is implemented in practice.

## Data Availability

All data produced in the present study are available upon reasonable request to the authors

## Contributors

SH and JC conceived the idea and developed the initial proposal with input from all co-authors. JC wrote the ethics applications, led the data collection, and contributed to data analysis and manuscript revision. AM led the data analysis and contributed to the manuscript draft, revision, and concepts. FK contributed to data collection, data analysis, manuscript draft and revision. AD and AC contributed to the data collection and manuscript revision. All authors contributed to manuscript development and approved the final manuscript.

## Data availability statement

All data relevant to the study are included in the article or uploaded as supplementary information.

## Acknowledgements

We thank members of our NIHR Patient and Public Involvement Project Advisory Board (Babatunde Tikare, Larysa Agbaso, Monika Hartmann, Saliha Majeed) and additional members of our Project Board for valuable contributions.

## References

1. Deal A, Halliday R, Crawshaw AF, Hayward SE, Burnard A, Rustage K, et al. Migration and outbreaks of vaccine-preventable disease in Europe: a systematic review. The Lancet Infectious Diseases. 2021.

2. Mipatrini D, Stefanelli P, Severoni S, Rezza G. Vaccinations in migrants and refugees: a challenge for European health systems. A systematic review of current scientific evidence. Pathogens and Global Health. 2017;111(2):59–68.

3. Jablonka A, Happle C, Grote U, Schleenvoigt BT, Hampel A, Dopfer C, et al. Measles, mumps, rubella, and varicella seroprevalence in refugees in Germany in 2015. Infection. 2016;44(6):781–7.

4. Norman FF, Comeche B, Martínez-Lacalzada M, Pérez-Molina J-A, Gullón B, Monge-Maillo B, et al. Seroprevalence of vaccine-preventable and non-vaccine-preventable infections in migrants in Spain. Journal of Travel Medicine. 2021;28(4).

5. Charania NA, Gaze N, Kung JY, Brooks S. Vaccine-preventable diseases and immunisation coverage among migrants and non-migrants worldwide: A scoping review of published literature, 2006 to 2016. Vaccine. 2019;37(20):2661–9.

6. Hargreaves S, Nellums LB, Ravensbergen SJ, Friedland JS, Stienstra Y, On Behalf Of The ESGITM Working Group On Vaccination In Migrants. Divergent approaches in the vaccination of recently arrived migrants to Europe: a survey of national experts from 32 countries, 2017. Eurosurveillance. 2018;23(41).

7. Crawshaw AF, et al. Defining the determinants of vaccine uptake and under-vaccination in migrant populations in Europe to improve routine and COVID-19 vaccine uptake: a systematic review. 2022 (Accepted, Lancet Infectious Diseases). https://www.medrxiv.org/content/10.1101/2021.11.08.21266058v1.

8. World Health Organization. Immunization Agenda 2030 2020 [updated 1 April 2020. Available from: https://www.who.int/teams/immunization-vaccines-and-biologicals/strategies/ia2030.

9. WHO. Leave no one behind. Guidance for planning and implementing catch-up vaccination. Geneva: WHO, 2021..

10. World Health Organization (WHO). Immunization Agenda 2030: A Global Strategy to Leave No One Behind. 2020.

11. Noori T, Hargreaves S, Greenaway C, van der Werf M, Driedger M, Morton RL, et al. Strengthening screening for infectious diseases and vaccination among migrants in Europe: What is needed to close the implementation gaps? Travel Med Infect Dis. 2021;39:101715.

12. European Centre for Disease Prevention and Control. Public health guidance on screening and vaccination of migrants in the EU/EEA. Available from: https://www.ecdc.europa.eu/en/publications-data/public-health-guidance-screening-and-vaccination-infectious-diseases-newly. ECDC. Sweden: Stockholm; 2018.

13. Public Health England. Vaccination of individuals with uncertain or incomplete immunisation status. Updated 26th August 2021. Available from: https://www.gov.uk/government/publications/vaccination-of-individuals-with-uncertain-or-incomplete-immunisation-status/vaccination-of-individuals-with-uncertain-or-incomplete-immunisation-status [Accessed 21st September 2021].

14. Morse JM (1995) The Significance of Saturation. Qual Health Res 5(2): 147–149 Available from: [https://journals.sagepub.com/doi/10.1177/104973239500500201].

15. Braun V, Clarke V. Thematic analysis. In: Cooper H, Camic PM, Long DL, editors. APA handbook of research methods in psychology, 2: research designs: quantitative, qualitative, neuropsychological, and biological. Washington, DC: American Psychological Association; 2012.

16. AuYOung M, et al. Addressing racial/ethnic inequities in vaccine hesitancy and uptake: lessons learned from the California alliance against COVID-19. J Behavioural Med 2022; 22nd Jan https://link.springer.com/article/10.1007/s10865-022-00284-8.

17. Socha A, Klein J. What are the challenges in the vaccination of migrants in Norway from healthcare provider perspectives? A qualitative, phenomenological study BMJ Open 2020;10:e040974. doi: 10.1136/bmjopen-2020-040974.

18. Moussaoui S, Aurousseau AM, Nappez S, Cornaglia J, Delobre G, Blanchi S, et al. Immunization Catch-Up for Newly Arrived Migrants in France: A Cross-Sectional Study among French General Practitioners. 2021;9(6):21.

19. Ravensbergen SJ, et al. National approaches to the vaccination of recently arrived migrants in Europe: a comparative policy analysis across 32 European countries. Trav Med and Infect Dis 2019; 33–38..

20. Giambi C, Del Manso M, Marchetti G, Olsson K, Adel Ali K, Declich S, et al. Immunisation of migrants in EU/EEA countries: Policies and practices. Vaccine. 2019;37(36):5439–51.

21. Evlampidou I, et al. Low hepatitis B testing among migrants: a cross-sectional study in a UK city. Br J Gen Pract 2016; 66)647): e382–e391..

22. Crawshaw AF, Deal A, Rustage K, Forster AS, Campos-Matos I, Vandrevala T, et al. What must be done to tackle vaccine hesitancy and barriers to COVID-19 vaccination in migrants? Journal of Travel Medicine. 2021;28(4):01.

23. Jama A, et al. Perspectives on the measles, mumps and rubella vaccination among Somali Mothers in Stockholm. Int J Environmental Research and Public Health 2018; 15: 11.https://www.mdpi.com/1660-4601/15/11/2428.

24. Kasstan B. Vaccines and vitriol: an anthropological commentary on vaccine hesitancy, decision-making and interventionism among religious minorities. Anthrop & Med 2020; 13 Nov: 411–419 https://www.tandfonline.com/doi/full/10.1080/13648470.2020.1825618?casa_token=DiweSjBRwMQAAAAA%3Aw1JFAzqfbT8KsQTnz72bbpGy7pcmgRjehDA8OeGAMmOD1esZFPHL0IWhxq2M-p0vU5Z4F3-cFEaZTQ.

25. Larson HJ, et al. Understanding vaccine hesitancy around vaccines and vaccination from a global perspective: a systematic review of published literature, 2007-2012. Vaccine; 32: 19: 2150–2159.

26. WHO. Immunization coverage. July 15, 2021. Available from: https://www.who.int/news-room/fact-sheets/detail/immunization-coverage.

27. Tankwanchi AS, Bowman B, Garrison M, Larson H, Wiysonge CS. Vaccine hesitancy in migrant communities: a rapid review of latest evidence. Curr Opin Immunol. 2021;71:62–8.

28. Doctors of the World UK. Vaccine Confidence Toolkit. England: London. 2021. [Available from: https://www.doctorsoftheworld.org.uk/what-we-stand-for/supporting-medics/vaccine-confidence-toolkit/

29. Hui, C., Dunn, J., Morton, R., Staub, L. P., Tran, A., Hargreaves, S., Greenaway, C., Biggs, B. A., Christensen, R., & Pottie, K. (2018). Interventions to Improve Vaccination Uptake and Cost Effectiveness of Vaccination Strategies in Newly Arrived Migrants in the EU/EEA: A Systematic Review. International journal of environmental research and public health, 15(10), 2065. https://doi.org/10.3390/ijerph15102065.

